# Brain natural frequencies as physiologically meaningful biomarkers for machine-learning detection of Parkinson’s disease

**DOI:** 10.64898/2025.12.09.25341882

**Authors:** Lydia Arana, Joachim Gross, Almudena Capilla

**Affiliations:** Departamento de Psicología Biológica y de la Salud, Facultad de Psicología, Universidad Autónoma de Madrid, Madrid, Spain; Institute for Biomagnetism and Biosignal Analysis, University of Münster, Münster, Germany

**Keywords:** Brain oscillations, Electroencephalography, Natural frequencies, Machine learning, Oscillopathies, Parkinsońs disease, Resting state

## Abstract

In this study, we investigated whether individual brain maps of natural frequencies derived from EEG can serve as physiologically meaningful biomarkers for Parkinson’s disease (PD), a disorder characterized by altered oscillatory activity. Data were gathered from three independent, openly available EEG databases. Natural frequency maps were extracted for 57 PD participants and 57 sex- and age-matched healthy controls (HC). The group-level brain maps showed region-specific slowing of oscillatory activity in PD, particularly in frontomedial and left frontolateral cortices, as well as hemispheric asymmetries consistent with the lateralized onset of motor symptoms. A subset of 100 participants (50 PD, 50 matched controls) was then used to train and internally validate three machine learning classifiers—support vector machine (SVM), random forest, and logistic regression—under different PCA-based dimensionality reduction and feature normalization schemes. Among all models, SVM achieved the best performance (AUC ≈ 77%). This classifier was subsequently retrained on the full set of 100 participants and evaluated on an independent hold-out test set of 14 participants, achieving an AUC of 75.5% with 71.4% accuracy. Feature contribution analysis highlighted the regions that were most informative for classification toward each class, complementing the group-level observations. Overall, these results indicate that natural frequency mapping captures disease-related alterations in cortical dynamics and provides interpretable features for EEG-based diagnostic tools, opening new avenues for biomarker development in oscillopathies.

## 1. Introduction

Or brains exhibit intrinsic rhythmic activity, characterized by frequency-specific patterns—its natural frequencies—that reflect the oscillatory dynamics of neuronal networks (Buzsáki, 2006; Capilla et al., 2022; Keitel & Gross, 2016). The brain’s natural frequencies can be captured using electro- or magnetoencephalography (EEG/MEG), either following external perturbations with transcranial magnetic stimulation (TMS) (Amengual et al., 2019; Ferrarelli et al., 2012; Rosanova et al., 2009) or directly from ongoing electrophysiological activity (Capilla et al., 2022; Frauscher et al., 2018; Kalamangalam et al., 2020; Keitel & Gross, 2016; Mahjoory et al., 2020; Mellem et al., 2017). Such spectral profiles have been found to be characteristic of specific brain areas, to the extent that they can be used to differentiate and classify regions based on their intrinsic oscillatory dynamics (Keitel & Gross, 2016; Komorowski et al., 2023). Moreover, natural frequencies can also be characterized at the individual level, acting as a neural fingerprint that remains stable over the years (Arana et al., 2025).

Oscillatory activity is essential for brain function and communication (Buzsáki & Watson, 2012; Fries, 2015; Varela et al., 2001) and its alteration is deeply related to a wide range of neurological and psychiatric disorders, also referred to as “oscillopathies” (Buzsáki et al., 2013). Among these, Parkinson’s disease (PD) provides a well-established example, where abnormal rhythmic patterns are associated with core motor and cognitive symptoms (Nimmrich et al., 2015; Schnitzler & Gross, 2005).

PD is a neurodegenerative disorder marked by the loss of dopaminergic neurons in the substantia nigra, clinically characterized by bradykinesia, muscular rigidity, tremor, and postural instability, as well as non-motor symptoms—including olfactory dysfunction, cognitive impairment, psychiatric manifestations, sleep disorders, and autonomic dysfunction—which may precede motor dysfunction by several years (Chaudhuri & Schapira, 2009; Kalia & Lang, 2015). Accurate diagnosis of PD is essential for both prognosis and therapeutic management. Although diagnostic precision has improved in recent years, it still relies primarily on clinical features and varies with disease stage and clinician expertise, typically reaching around 80% (Rizzo et al., 2016; Virameteekul et al., 2023).

Alterations in brain oscillations are commonly observed in patients with PD (Nimmrich et al., 2015). While findings show some variability across studies and frequency bands, most evidence points to a reduction in alpha (8–13 Hz) and beta (13–30 Hz) power together with an increase in delta (0.5–4 Hz) and theta (4–8 Hz) power in resting state (Bosboom et al., 2006; Kotini et al., 2005; Morita et al., 2009; Serizawa et al., 2008; Soikkeli et al., 1991; Stoffers et al., 2007; Zhao et al., 2025); a constellation of changes generally described as “slowing” of EEG/MEG activity (Geraedts et al., 2018). However, although these studies refer to slowing, most focus only on band power amplitudes rather than on peak frequency values. Only a few studies have assessed peak frequencies (Kotini et al., 2005; Soikkeli et al., 1991; Zhao et al., 2025), particularly the alpha peak, which is shifted toward lower frequencies compared to healthy controls (HC). While this provides direct evidence of EEG/MEG slowing, it remains limited to the alpha band. We hypothesize that assessing exact frequency values throughout different cortical regions could contribute to extend these findings.

Recent efforts to identify characteristic oscillatory signatures to support PD diagnosis have been boosted by the exponential growth of machine learning tools. Among the available techniques, EEG stands out for its accessibility and cost-effectiveness, making EEG-based approaches particularly promising for this purpose (Maitín et al., 2020). However, there is substantial variability between studies, and those achieving the highest accuracies (>85%) distinguishing PD from HC should be interpreted with caution, as they may reflect certain methodological considerations, such as relatively small patient samples (Aljalal et al., 2022; Anjum et al., 2020; Vanneste et al., 2018; Waninger et al., 2020; Yuvaraj et al., 2018), non-standardized preprocessing protocols (Aljalal et al., 2022; Vanneste et al., 2018; Waninger et al., 2020; Yuvaraj et al., 2018), and exclusive reliance on internal cross-validation (Aljalal et al., 2022; Vanneste et al., 2018; Yuvaraj et al., 2018).

In the present study, we aimed to evaluate the brain’s natural frequencies as potential biomarkers of PD by testing a binary classifier distinguishing PD patients from HC. Natural frequencies represent simple yet informative features that can be reliably extracted from resting-state EEG/MEG recordings, complementing traditional oscillatory power measures. To overcome potential methodological constraints of earlier studies, we employed a relatively large sample of sex- and age-matched PD patients and HC, a fully standardized and reproducible analysis pipeline, and an independent hold-out test set, enabling external validation and enhancing the generalizability of our results.

## 2. Materials and methods

### 2.1 Participants

We used data from three openly available databases: the Two Decades-Brainclinics Research Archive for Insights in Neurophysiology (TDBRAIN) database (Van Dijk et al., 2022), the Linear Predictive Coding for PD at Rest (PD LPC Rest) dataset (Anjum et al., 2020), and the Latin American Brain Health Institute (BrainLat) dataset (Prado et al., 2023).

We selected participants from these databases to obtain a sample as homogeneous as possible of PD patients and sex- and age-matched HC, for a total of 114 individuals (57 PD and 57 HC). For classifier evaluation, we randomly split the sample into a training/validation set of 100 participants (87.7% of the total; 50 PD and 50 HC) and a hold-out test set of 14 participants (12.3% of the total; 7 PD and 7 HC), while maintaining the original sex/age matching within each group.

#### Training/validation set

Within the training/validation set (N=100), the mean age of the PD patients was 66.4 ± 9.6 years [M ± SD], ranging from 43 to 84 years, and included 18 females and 32 males. The mean age of the HC participants was 66.3 ± 10.9 years [M ± SD], ranging from 40 to 84 years, and also included 18 females and 32 males. There was no significant difference in age between PD patients and their sex- and age-matched HC (*t*_(98)_ = 0.08, *p* = 0.94, Welch two-sample t-test). The mean age difference between PD patients and their sex- and age-matched HC was 1.56 ± 1.58 years [M ± SD], with a maximum difference of 6 years.

Among the PD patients, 12 were obtained from TDBRAIN, 25 from PD LPC Rest, and 13 from the BrainLat dataset. Among the HC participants, 9 were obtained from TDBRAIN, 23 from PD LPC Rest, and 18 from the BrainLat dataset.

#### Hold-out test set

Within the hold-out test set (N=14), the mean age of the PD patients was 65.4 ± 8.7 years [M ± SD], ranging from 51 to 75 years, and included 4 females and 3 males. The mean age of the HC participants was 65.1 ± 9.8 years [M ± SD], ranging from 48 to 75 years, and also included 4 females and 3 males. There was no significant difference in age between PD and HC (*t*_(12)_ = 0.06, *p* = 0.95, Welch two-sample t-test). The age difference between PD patients and their sex- and age-matched HC was 1.1 ± 1.2 years [M ± SD], with a maximum difference of 3 years.

Among the PD patients, 2 were obtained from TDBRAIN, 2 from PD LPC Rest, and 3 from the BrainLat dataset. Among the HC participants, 4 were obtained from PD LPC Rest and 3 from the BrainLat dataset.

### 2.2 Databases

#### TDBRAIN

The Two Decades-Brainclinics Research Archive for Insights in Neurophysiology (TDBRAIN; Van Dijk et al., 2022) is a clinical EEG lifespan database comprising raw resting-state EEG recordings complemented with clinical and demographic information. The dataset includes a heterogeneous sample of 1274 individuals—patients with various neuropsychiatric disorders as well as healthy volunteers—collected between 2001 and 2021 as part of routine clinical care and applied neuroscience projects at the Research Institute Brainclinics (Brainclinics Foundation, Nijmegen, The Netherlands). All participants provided informed consent, explicitly agreeing to have their data included in the database for open access.

We selected the patients with a formal diagnosis of PD and controls with confirmed healthy status, with the information about age and sex as provided by the dataset. For these participants, behavioral measures are also included for an auditory oddball task performed after the resting-state EEG, as well as information on smoking, alcohol use, drug consumption, years of education, and hours of sleep. However, the dataset does not provide information on the clinical characterization of PD patients, such as disease stage, motor symptoms, or cognitive status.

#### PD LPC Rest

The PD Linear Predictive Coding at Rest dataset (PD LPC Rest; Anjum et al., 2020) was compiled for PD research between the University of New Mexico (UNM, Albuquerque, New Mexico) and the University of Iowa (Iowa City, Iowa), including a sample of 82 PD patients and healthy participants from New Mexico (54 individuals) and Iowa (28 individuals). All participants provided written informed consent and received compensation for their participation.

Demographic information on participants’ age, sex, and years of education is included in the dataset, along with extensive clinical and neuropsychological evaluations.

#### The BrainLat dataset

The Latin American Brain Health Institute (BrainLat; Prado et al., 2023) released a multimodal neuroimaging dataset of 780 participants from Latin America—patients with various neurodegenerative diseases as well as healthy volunteers—collected through a multicentric effort across five countries. All participants provided written consent, explicitly authorizing open publication of anonymized data.

The EEG data for PD patients and healthy participants were collected at the Neurology Department, Geroscience Center for Brain Health and Metabolism (Santiago; 45 participants) and the *Centro de Neurociencia Cognitiva, Universidad de San Andrés* (Buenos Aires; 25 participants). Demographic information is available for these participants, including sex, age, years of education, and laterality. In addition, the dataset includes extensive cognitive and behavioral assessments spanning multiple modalities, such as visual, verbal, and motor performance, emotion recognition, and general cognitive status.

### 2.3 EEG acquisition

#### TDBRAIN

EEG was acquired using a 26-channel Compumedics Quickcap or ANT-Neuro Waveguard Cap with sintered Ag/AgCl electrodes, grounded at AFz, sampled at 500 Hz, and low-pass filtered at 100 Hz. Skin impedance was kept below 10 kΩ using a conductive, non-toxic aqueous gel (Quick-Gel, Compumedics NeuroMedical Supplies, USA, or OneStep Cleargel).

Brain activity was recorded during resting state, which consisted of a 2-minute Eyes Open (EO) task, during which participants were asked to rest quietly with their eyes open while focusing on a red dot at the center of a computer screen, and a 2-minute Eyes Closed (EC) task, in which participants were asked to close their eyes while maintaining the same posture as in the EO task. Only the Eyes Closed (EC) recordings were used in this study to align with the BrainLat’s eyes-closed procedure.

#### PD LPC Rest

EEG was recorded using a 64-channel Brain Vision system with sintered Ag/AgCl electrodes, sampled at 500 Hz and band-pass filtered between 0.1 and 100 Hz, with the reference set to CPz (UNM) and Pz (Iowa).

Brain activity was recorded during resting state, comprising EC and EO conditions in the UNM dataset and only the EO condition in the Iowa dataset. Only the UNM-EC data were used in this study to align with BrainLat’s eyes-closed procedure.

#### The BrainLat dataset

EEG was recorded using a 128-channel BioSemi ActiveTwo system with sintered Ag/AgCl electrodes, band-pass filtered between 0.03 and 100 Hz, and referenced to the common mode sense (CMS) electrode with a driven right leg (DRL) ground. The authors did not report the acquisition sampling rate; however, they stated that the data were downsampled to 512 Hz, which corresponds to the sampling rate of the shared dataset.

Brain activity was recorded during a 10-minute EC resting-state session, while participants sat in a dimly lit, sound-attenuated, and electromagnetically shielded room.

### 2.4 Preprocessing

The scripts necessary to reproduce all the analysis and figures are available at https://github.com/necog-UAM. Analyses were carried out using FieldTrip (version 20230118; Oostenveld et al., 2011), EEGLAB (version 2023.0; Delorme & Makeig, 2004) and in-house Matlab code (R2025a version). For data preprocessing, we adapted the automatic pipeline DISCOVER-EEG (Gil Ávila et al., 2023) (see Figure 1 for a schematic overview of the methodological procedure).

**Figure 1.**
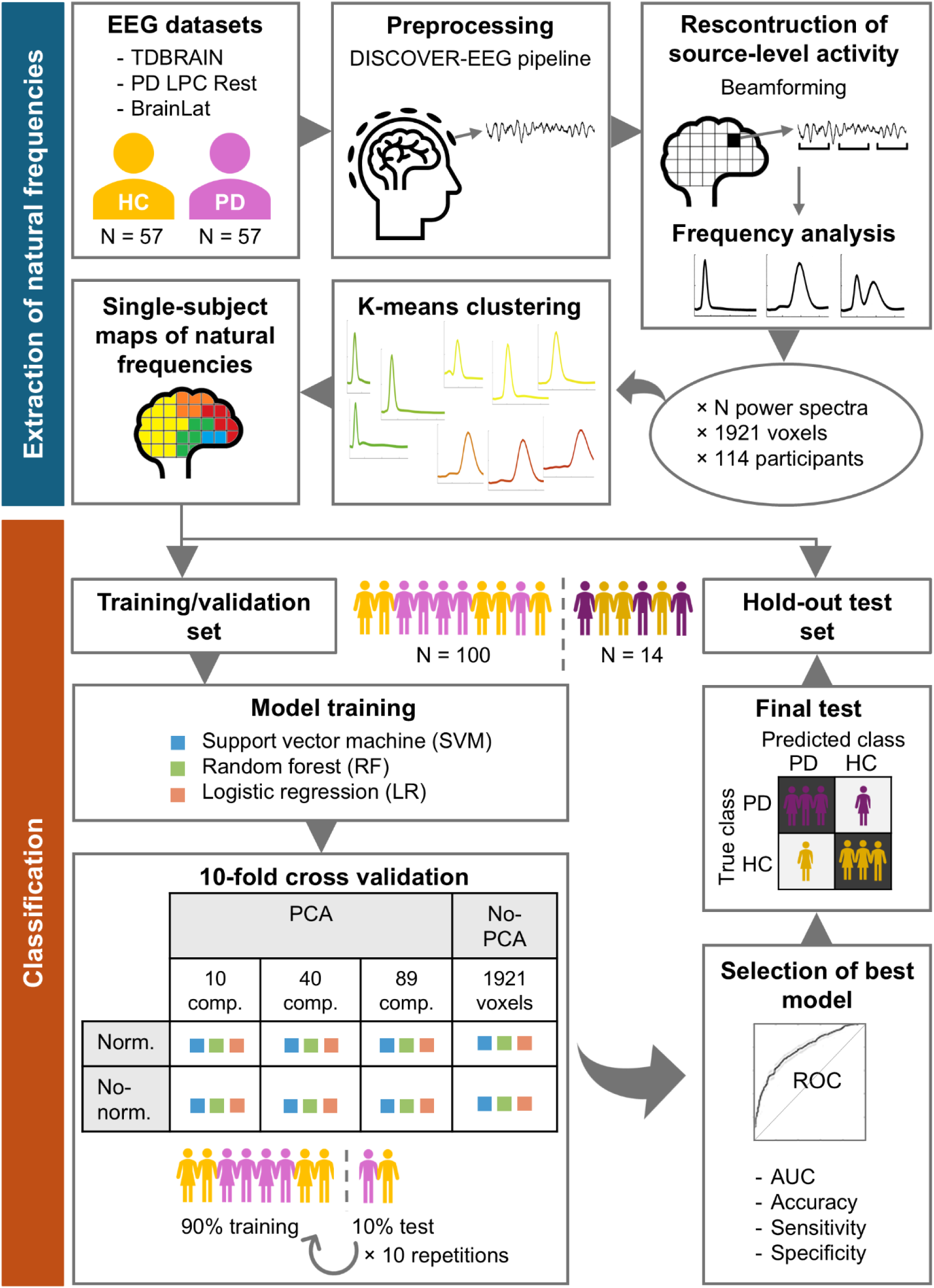
Overview of the methodological procedure. Pipeline used for the extraction and classification of individuals based on their brain’s natural frequencies. Abbreviations: HC: healthy controls; PD: Parkinson’s disease; PCA: principal component analysis; comp.: components; norm.: normalization; ROC: receiver operating characteristic; AUC: area under the ROC curve.

For the BrainLat dataset, EEG signals had already been preprocessed by the dataset authors prior to data sharing. According to the description provided in Prado et al. (2023), preprocessing of the BrainLat dataset consisted of re-referencing to the average, applying a 0.5–40 Hz band-pass filter using an 8th-order zero-phase Butterworth, performing Independent Component Analysis (ICA) to correct blinks and eye movements, and identifying malfunctioning channels using a semiautomatic detection method, which were then replaced using weighted spherical interpolation. Therefore, this dataset was excluded from the corresponding preprocessing steps of the pipeline described below, except for the detection of bad segments. We also included resampling to 500 Hz to match the sampling rate of the other datasets and visually inspected the provided data to ensure its quality.

Power line artifacts were reduced by means of spectrum interpolation (Leske & Dalal, 2019) at 50/100/150 Hz for the TDBRAIN dataset and at 60/120/180 Hz for the PD LPC Rest dataset. Data were then high-pass filtered with a default transition band of 0.25–0.75 Hz. Channels were marked as artifactual and discarded if (1) they remained flat for more than 5 seconds, (2) their z-scored noise-to-signal ratio exceeded the default threshold of 4, or (3) their correlation with neighboring channels was below 0.8. These parameters follow the defaults described in Pernet et al. (2021) and Gil Ávila et al. (2023). The data were then re-referenced to the average reference.

Next, five repetitions of the following sequence were applied. (1) Artifact components were removed using ICA with the ‘runica’ algorithm, which automatically identifies and categorizes components (Pion-Tonachini et al., 2019). Components with a probability greater than 80% of being classified as ‘Muscle’ or ‘Eye’ were subtracted (Gil Ávila et al., 2023; Pernet et al., 2021). (2) Channels previously excluded were interpolated with spherical splines (Perrin et al., 1989). (3) Time periods containing artifacts were annotated but not eliminated, to avoid edge effects in the continuous data. In the original DISCOVER-EEG pipeline (Gil Ávila et al., 2023), such segments are rejected using Artifact Subspace Reconstruction (ASR; Mullen et al., 2015), which detects intervals with abnormally high power. In our implementation, the raw data were retained prior to segment rejection, and the artifactual intervals were instead marked for exclusion from subsequent analyses. The parameters used to define bad segments were adjusted relative to those of Pernet et al. (2021) and Gil Ávila et al. (2023). In our case, segments with variance 60 times greater than calibration data were marked as bad, compared to the default parameter of 20, which tended to eliminate alpha activity. Segments containing ≥10% of bad channels were also flagged—a stricter criterion than the default 25%—to ensure more reliable artifact detection. The tolerance threshold for standard deviations was set to 9.

Among the five repetitions, the cleaned dataset exhibiting the bad-segment mask closest to the average across all iterations was selected as the final output. The number of interpolated bad channels was 1.8 ± 1.2 for TDBRAIN and 3.5 ± 2.4 for PD LPC Rest. The number of rejected eye components was 0.9 ± 0.8 for TDBRAIN and 2.8 ± 1.0 for PD LPC Rest. The number of rejected muscle components was 0.3 ± 0.6 for TDBRAIN and 6.4 ± 4.4 for PD LPC Rest. Finally, prior to source reconstruction, clean data were visually inspected to confirm the effectiveness of the preprocessing procedure.

### 2.5 Reconstruction of source-level activity

The pipeline for source reconstruction was adapted to EEG from the original detailed in Capilla et al. (2022) for MEG data.

To obtain the head model, the standard MRI (Holmes, 1998) was co-registered to the EEG standard 10–5 coordinate system (Oostenveld et al., 2003; Oostenveld & Praamstra, 2001), selecting the corresponding EEG channels in our data. Only channels common to all three datasets were retained, leaving 25 channels for further source reconstruction and analysis. This selection was necessary to homogenize localization accuracy when using low-density EEG—as in the case of TDBRAIN (26 channels)—since a higher number of electrodes (BrainLat and PD LPC, 128 and 64 channels, respectively) reduces localization error (Song et al., 2015). Retaining all available electrodes would have reduced the comparability of source-reconstructed activity across the three datasets.

The forward model was computed using a realistic single shell volume conductor model (Nolte, 2003). A standard 3D-grid with 1-cm resolution was adapted to the head model brain volume and lead fields were computed for each grid point (i.e., voxel). Voxels located outside the cerebral cortex or the hippocampus (i.e., cerebellum and subcortical structures) were excluded, leaving 1925 voxels for further analysis. Four dipoles overlapped with electrode locations and were therefore discarded, yielding a final reconstruction for 1921 voxels. We then employed linearly constrained minimum variance (LCMV) beamforming (Van Veen et al., 1997) to estimate source-level activity. The spatial filter weights were derived from the covariance of the clean data, using a regularization parameter lambda of 10%. These beamforming weights were then used to reconstruct source-space time series from the sensor-level data. Finally, to account for the beamformer’s bias toward the center of the head (Shapira Lots et al., 2016), we normalized the source-level data of each voxel by dividing it by its standard deviation across time.

### 2.6 Frequency analysis on source-level data

Frequency analysis was performed identically for all three datasets on the reconstructed signal. Since the duration of the recordings differed substantially across participants and datasets (TDBRAIN ∼2 minutes, PD LPC ∼2–10 minutes, BrainLat ∼10 minutes), we used the full length of each recording to maximize the number of spectra per participant and thereby improve the extraction of single-subject natural frequency maps (Arana et al., 2025).

A Hanning-tapered sliding window Fourier transform was applied to the source-reconstructed signals, using 200-ms steps (Arana et al., 2025). Power was calculated across 61 logarithmically spaced frequency bins, ranging from 1.7 to 34.8 Hz, as earlier findings indicated that higher frequencies primarily reflected artifactual activity at rest (Capilla et al., 2022). The sliding window’s width was adapted to five cycles per frequency bin, to attenuate the 1/f aperiodic component (Capilla et al., 2022). To prevent edge effects, 5.8-second time intervals (corresponding to 10 cycles at the lowest frequency of 1.7 Hz) at the start and end of each artifact-free segment were excluded from the analysis. Thus, we obtained a set of 1281.6 ± 913.5 available power spectra per voxel and participant.

### 2.7 Cluster analysis of power spectra

To identify patterns of source-reconstructed oscillatory activity, k-means clustering was performed using the cosine distance metric to maximize differences between clusters based on the shape of the power spectra (Keitel & Gross, 2016). From the pool of all available spectra, 300 power spectra were randomly selected per voxel and participant. This number corresponded to the participant with the lowest number of available spectra. Thus, a total of 114 participants × 1921 voxels × 300 power spectra were included into the clustering. The number of clusters was set to 25, as the algorithm’s performance has been shown to plateau at this number for this type of data (Capilla et al., 2022). Five clustering replicates were run with a maximum of 200 iterations each. We retained the solution with the lowest within-cluster sum of distances.

### 2.8 Single-subject brain maps of natural frequencies

After training the k-means clustering model, the single-subject maps of natural frequencies were computed. For each participant, all power spectra previously obtained for each voxel (see Section 2.6) were assigned to the cluster whose centroid showed the smallest cosine distance (Keitel & Gross, 2016).

The oscillatory frequency characterizing each cluster was then determined by identifying local maxima in the centroid power spectra. Centroids without identifiable peaks were excluded, as the presence of a spectral peak is necessary to confirm genuine oscillatory activity (Donoghue et al., 2022). When two peaks were detected, a check for harmonic relationships (e.g., 10 and 20 Hz, within a ±1 Hz tolerance) was performed. In cases of harmonicity, only the fundamental frequency was retained. If no harmonic relationship was found (e.g., 7 and 22 Hz), both frequencies were kept and assigned equal weights of 50% (Arana et al., 2025).

We next calculated the proportion of power spectra assigned to each cluster and normalized these proportions across voxels using z-scores. This procedure produced, for every participant and voxel, a z-score indicating the relative contribution of each cluster’s peak frequency. To avoid limitations imposed by the discrete centroid frequencies, the relative contribution of each cluster to each voxel was interpolated tenfold, yielding a finer spectral resolution.

Since sharp transitions may occur between the natural frequencies of neighboring voxels (e.g., delta vs. high-beta in frontal regions; Capilla et al., 2022), conventional spatial smoothing is not suitable for these data. Instead, we adopted the approach described in Arana et al. (2025). For each voxel, neighbors within a 1.5-cm radius were identified, and their z-values indicating the relative presence of each oscillatory frequency were collected, including that of the central voxel. One-sample t-tests against zero were then performed for each frequency bin to test whether a given frequency was consistently expressed in the local neighborhood, under the null hypothesis that the mean z-score across voxels was ≤ 0. The frequency with the highest t-value was assigned as the voxel’s natural frequency, as it represented the most consistent oscillatory component within the 1.5-cm neighborhood. Voxels where no frequency passed the significance threshold (p > 0.05) were assigned missing values. This procedure yielded, for each participant, a vector of 1921 frequency values, i.e., the single-subject natural frequency map, with each value corresponding to one voxel.

### 2.9 Group-level brain maps of natural frequencies

For visualization purposes, we generated the group-level maps for PD and HC, by aggregating the single-subject maps. For each voxel and clinical condition, we calculated the distribution of natural frequencies across participants. These distributions were represented as smoothed histograms using the 61 logarithmically spaced frequency bins (from 1.7 to 34.5 Hz) to ensure appropriate resolution across the entire frequency range. The most prominent peaks in each distribution were selected as candidates for group-level natural frequencies. Then, a Gaussian fit of all candidate peak frequencies was conducted, and the value shared by most participants was set as the natural frequency of the voxel in the group-level map.

### 2.10 Classifier fitting, feature processing, and performance evaluation in the training/validation set

We tested three commonly used classifiers—support vector machine (SVM), random forest (RF), and logistic regression (LR)—using the natural frequencies per voxel and participant as features (i.e., 1921 voxels and 100 participants) under four processing schemes, combining normalization and dimensionality reduction with principal component analysis (PCA; Jolliffe & Cadima, 2016): (1) no-normalization/no-PCA, (2) normalization-only/no-PCA, (3) PCA-only/no-normalization, and (4) both normalization and PCA.

SVM is a binary classifier that identifies a hyperplane separating two classes in a high-dimensional feature space, aiming to maximize the margin between them (Vapnik, 1999). A linear kernel SVM with L2 regularization was selected, as it is well-suited for high-dimensional data (Huang & Lin, 2016; Janecek et al., 2008). We used MATLAB’s `fitcsvm` function, with the regularization parameter set to 1 to balance margin maximization and classification errors.

RF is an ensemble classifier that constructs multiple decision trees on bootstrapped samples of the training data, and aggregates their predictions to improve robustness and reduce overfitting (Breiman, 2001). In this study, 100 trees were built using MATLAB’s `TreeBagger` function that generally provides stable ensemble performance without excessive computational cost.

LR is a linear classifier that estimates the probability of each class using a logistic function applied to a linear combination of the input features (Hosmer et al., 2000). In this study, we used LR with ridge regularization to prevent overfitting using MATLAB’s `fitclinear` function.

For feature processing schemes including normalization (normalization-only/no-PCA and both normalization and PCA), features in the training data of each fold were standardized using z-score normalization (subtracting the mean and dividing by the standard deviation computed from the training data), and the same parameters were applied to the corresponding test data to avoid data leakage in the cross-validation. In the case of both normalization and PCA, normalization was applied previous to dimensionality reduction.

For processing schemes including PCA-based dimensionality reduction (PCA-only/no-normalization and both normalization and PCA), PCA was performed on each fold using only the training data. Then, both training and test data were projected onto the resulting component space to prevent data leakage. Three levels of dimensionality reduction were tested—10, 40, and 89 components—representing high, medium, and low levels of feature compression. The upper limit of 89 components corresponds to the maximum number of valid principal components that can be computed within each training fold for the different processing schemes, given that 90 out of 100 participants were available for training (after 10-fold cross-validation) and data were mean-centered during normalization (i.e., n_train − 1 degrees of freedom).

For processing schemes without dimensionality reduction (no-normalization/no-PCA and normalization-only/no-PCA), the input features corresponded directly to the natural frequency values of each of the 1921 voxels.

Classifier performance was first assessed in the training/validation set for all classifiers and feature processing schemes using 10-repetition, 10-fold stratified cross-validation. In each fold, the model was trained on 9/10 of the data and evaluated on the remaining 1/10-fold, which was never used for training. Accuracy, sensitivity, and specificity were computed for each fold and repetition, as follows:

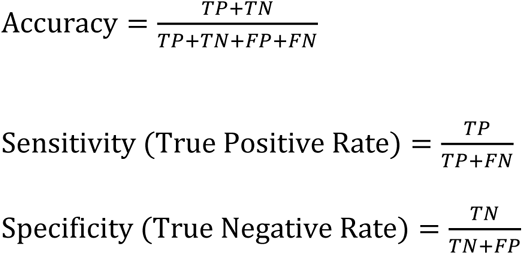

where *TP*, *TN*, *FP*, and *FN* denote true positives, true negatives, false positives, and false negatives, respectively.

We computed the mean across all 10 folds × 10 repetitions. Bootstrap resampling (1000 iterations) was applied to estimate 95% confidence intervals (CI) for all metrics, as the interval between the 2.5th and 97.5th percentiles of the distribution. The receiver operating characteristic (ROC) curve was also computed for each fold and repetition by plotting the true positive rate (𝑇𝑃𝑅 = 𝑇𝑃/(𝑇𝑃 + 𝐹𝑁)) against the false positive rate (𝐹𝑃𝑅 = 𝐹𝑃/(𝐹𝑃 + 𝑇𝑁)) across all decision thresholds (i.e., all possible probability cut-off values that uses the ROC algorithm. The corresponding area under the curve (AUC) was used as a threshold-independent measure of classification performance, with higher values indicating better discrimination between classes. We also calculated the mean AUC across the 10 folds × 10 repetitions along with its 95% CI using bootstrap resampling (1000 iterations).

### 2.11 Final model evaluation on hold-out test set

The optimal classification model and feature processing scheme were determined based on their performance across classification metrics (AUC, accuracy, sensitivity, and specificity). The selected classifier was trained once with all data from the training/validation set. The resulting final classification model was subsequently applied to the hold-out test set. AUC, accuracy, sensitivity, specificity, and the confusion matrix were obtained to evaluate classification performance on the hold-out test set.

### 2.12 Feature contributions

After training the selected classifier on the entire training/validation set for the final model evaluation, the contribution of each feature to the model’s decision was assessed using the Haufe transformation (Haufe et al., 2014). Specifically, the weight vector 𝑤 of the trained model in the original feature space was transformed using the covariance of the input features 𝑋, as follows:

This transformation maps the model weights and corrects for correlations between features, producing an interpretable pattern in the original feature space. The resulting values indicate how much each voxel contributes to the model’s prediction on the hold-out test set, allowing a meaningful interpretation of the model’s decision.

## 3. Results

### 3.1 Natural frequencies in PD and healthy individuals

Figure 2 shows the group-level maps of natural frequencies in healthy controls (HC, N = 57) and Parkinson’s disease patients (PD, N = 57). Both groups exhibited a similar pattern of natural frequency distribution. Higher frequencies within the beta band were predominantly observed in lateral and medial frontal regions, while lower frequencies in the delta and theta ranges were observed in medial prefrontal and temporal regions. Frequencies in the 7–13 Hz range (i.e., the alpha band) were prominently distributed over parieto-occipital regions. On a qualitative level, PD patients appeared to show a relative shift toward lower frequency values in several cortical regions, particularly in frontomedial areas, compared with HC. An asymmetry was also observed between the left and right frontolateral cortices, with greater slowing in the left hemisphere.

**Figure 2.**
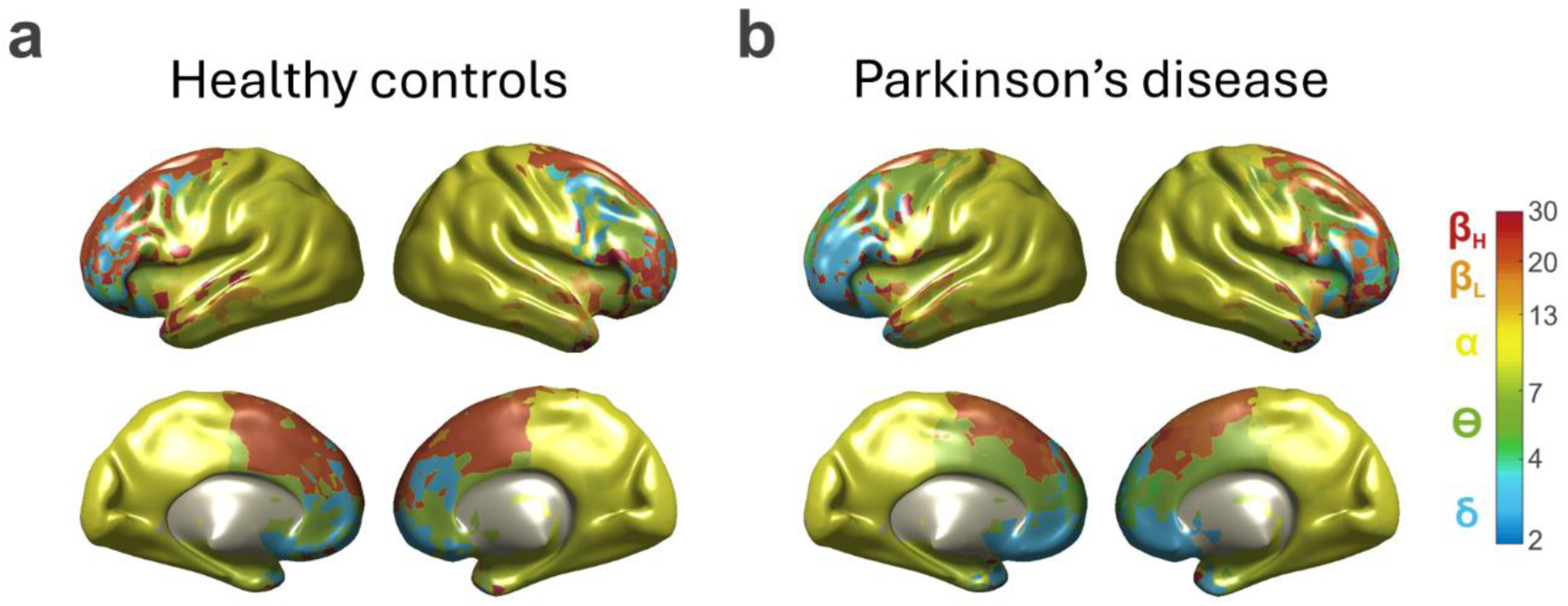
Voxel-wise distribution of natural frequencies in PD and healthy individuals. **(a)** Group-level map of natural frequencies in healthy controls (HC) (N = 57). **(b)** Group-level map of natural frequencies in Parkinson’s disease patients (PD) (N = 57). Each color corresponds to different oscillatory frequencies, as indicated in the colorbar on the right (delta: 2–4 Hz, theta: 4–7 Hz, alpha: 7–13 Hz, low beta: 13–20 Hz, high beta: 20–30 Hz).

### 3.2 Classifier performance across models and feature processing schemes

We tested three classifiers—support vector machine (SVM), random forest (RF), and logistic regression (LR)—under different processing schemes combining normalization and feature reduction via PCA (10, 40, and 89 components as features or, alternatively, 1921 voxels as features when dimensionality reduction was not applied).

Table 1 presents the results of all metrics (AUC, accuracy, sensitivity, and specificity) for each model across the different feature processing schemes. Figure 3 illustrates the ROC curves and accuracy obtained during cross-validation for each classification model. Mean accuracies across the 10 folds × 10 repetitions exceeded 50% for all models.

**Table 1.**
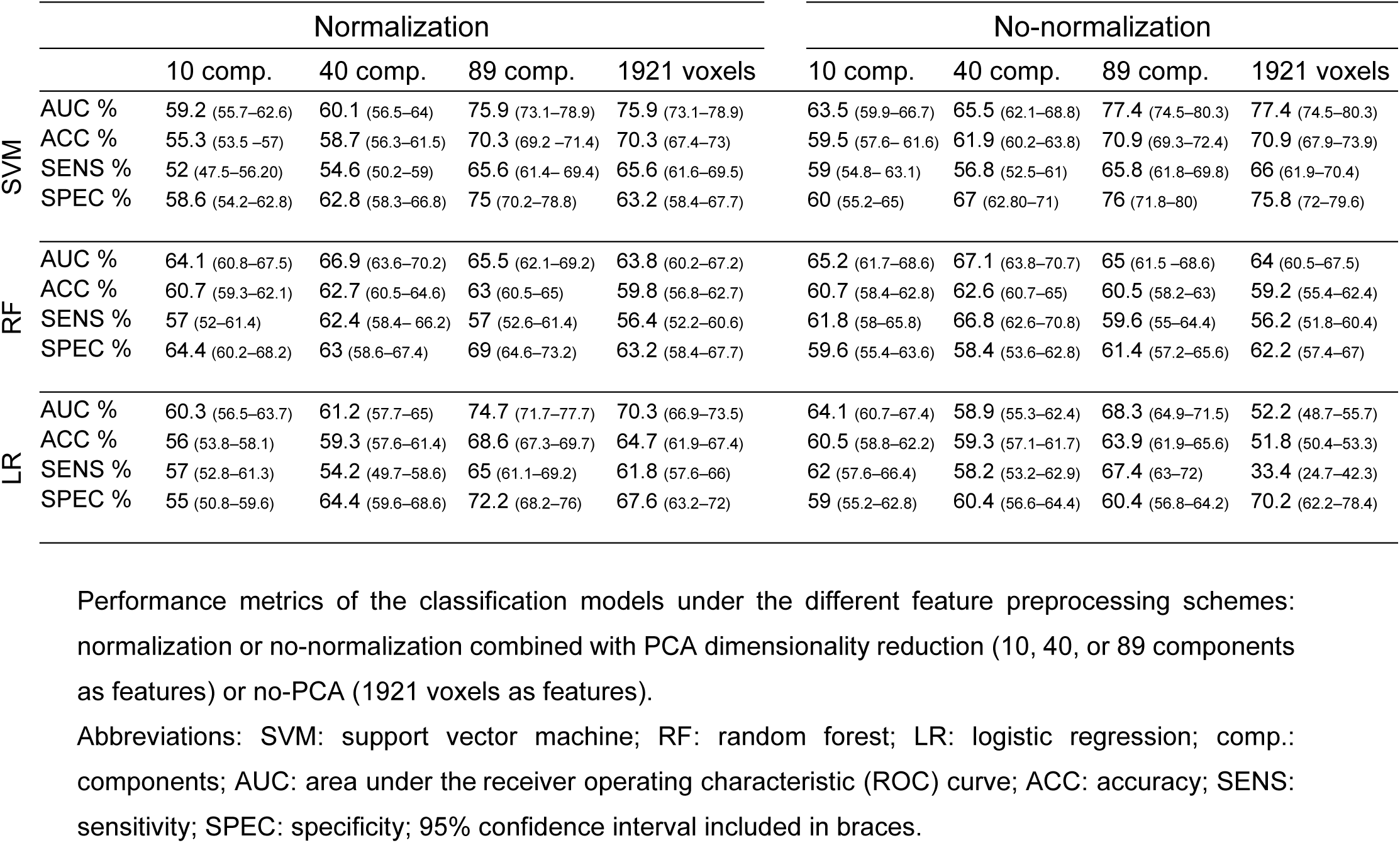
Performance metrics for the different classification models.

**Figure 3.**
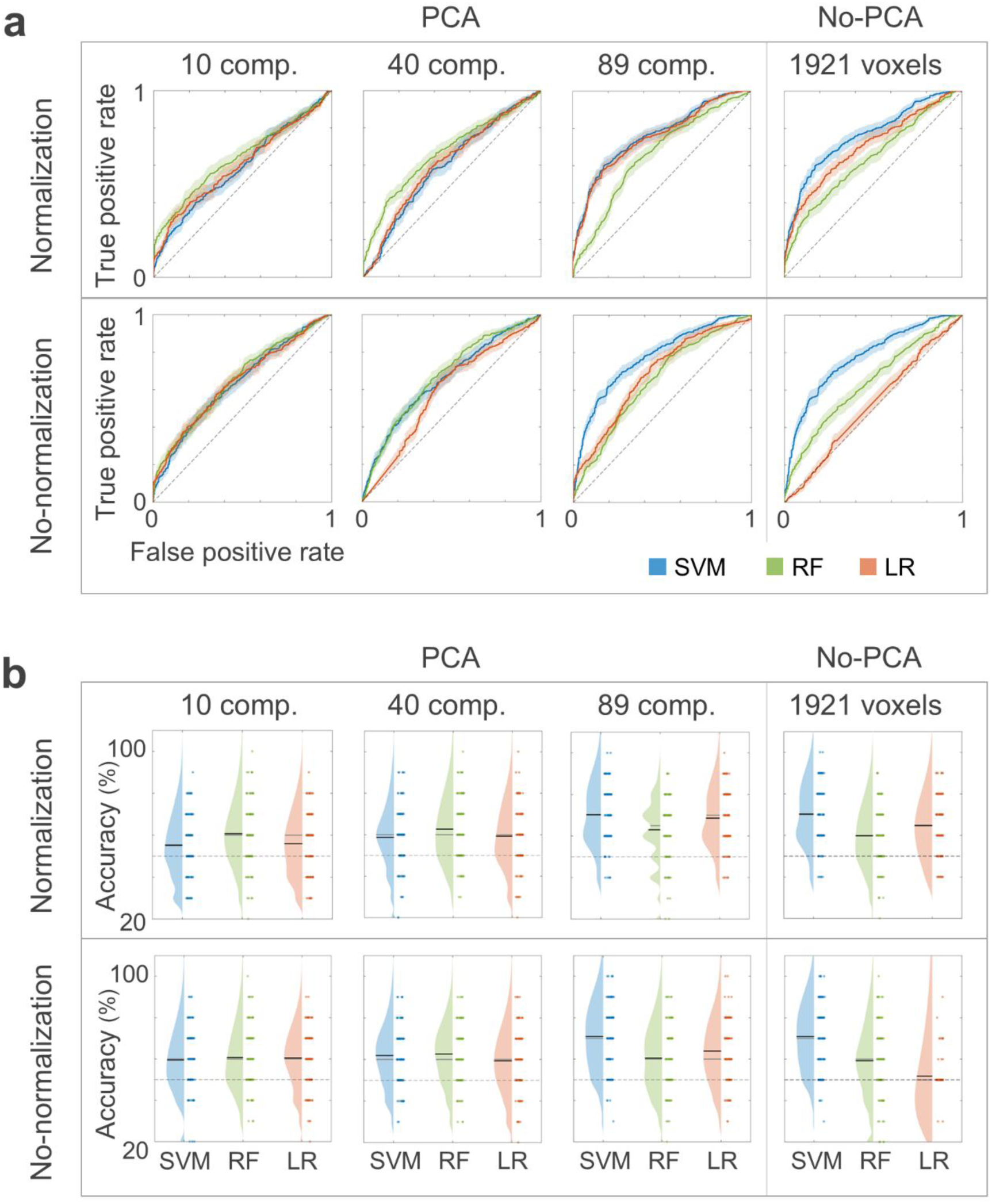
Summary of the performance of the classification methods under different feature preprocessing schemes across 10 folds × 10 repetitions. **(a)** Receiver operating characteristic (ROC) curves of support vector machine (SVM; blue), random forest (RF; green), and logistic regression (LR; orange) for different preprocessing schemes (normalization/no-normalization; PCA/no-PCA). Lighter colors indicate 95% confidence intervals. **(b)** Accuracy of SVM (blue), RF (green), and LR (orange) classifiers for different preprocessing schemes. Abreviations: comp. = components.

The ROC curves for each model across all 10 folds × 10 repetitions showed a consistent separation from the diagonal, indicating performance above chance levels, except for LR under the no-normalization/no-PCA scheme. The curves for SVM were generally closer to the upper-left corner of the plot, reflecting higher rate of true positives across thresholds compared to the RF and LR models (Figure 3a).

For SVM, performance improved both when using a higher number of components and when using all voxels as features and was highest when no normalization was applied: PCA (89 components), AUC = 77.4 (74.5–80.3), and no-PCA, AUC = 77.4 (74.5–80.3).

For RF, the best performance was achieved when using a moderate number of PCA components (40 components) either with normalization, AUC = 67.1 (63.8–70.7), or without normalization, AUC = 66.9 (63.6–70.2). Finally, LR performed better under normalization schemes, both when using a high number of components (89 components), AUC = 74.7 (71.7–77.7), and when PCA was not applied, AUC = 70.3 (66.9–73.5). Amongst the three models evaluated, SVM was the one achieving the highest scores, specifically under the no-normalization scheme for both 89 components and 1921 voxels. In both cases AUC was 77.4 (74.5–80.3), while accuracy was 70.9 (69.3–72.4) for the PCA reduction to 89 components and 70.9 (67.9–73.9) when PCA was not applied (Figure 3b).

### 3.3 Final model evaluation on hold-out test set and feature contributions

The classification model selected for the final evaluation was a SVM trained under the no-normalization/no-PCA scheme, as it achieved the best overall performance across all classification metrics (AUC, accuracy, sensitivity, and specificity). Although the no-normalization/PCA-only scheme with 89 components yielded comparable results, we opted for the most direct approach to enhance reproducibility and interpretability. Using the original natural frequency values per voxel proved sufficiently informative, avoiding additional transformations that might obscure the neurophysiological meaning of the features or complicate replication across datasets (Woo et al., 2017).

The final SVM model was trained on the complete training/validation set (N = 100) and tested on the independent hold-out test set (N = 14), which was never used for model selection, cross-validation, or training. The performance of the final model is shown in Figure 4. The model achieved an AUC of 75.5%, with an overall accuracy of 71.4%. Both sensitivity and specificity were 71.4%, indicating balanced performance across classes. In total, 10 participants were correctly classified and 4 were misclassified.

**Figure 4.**
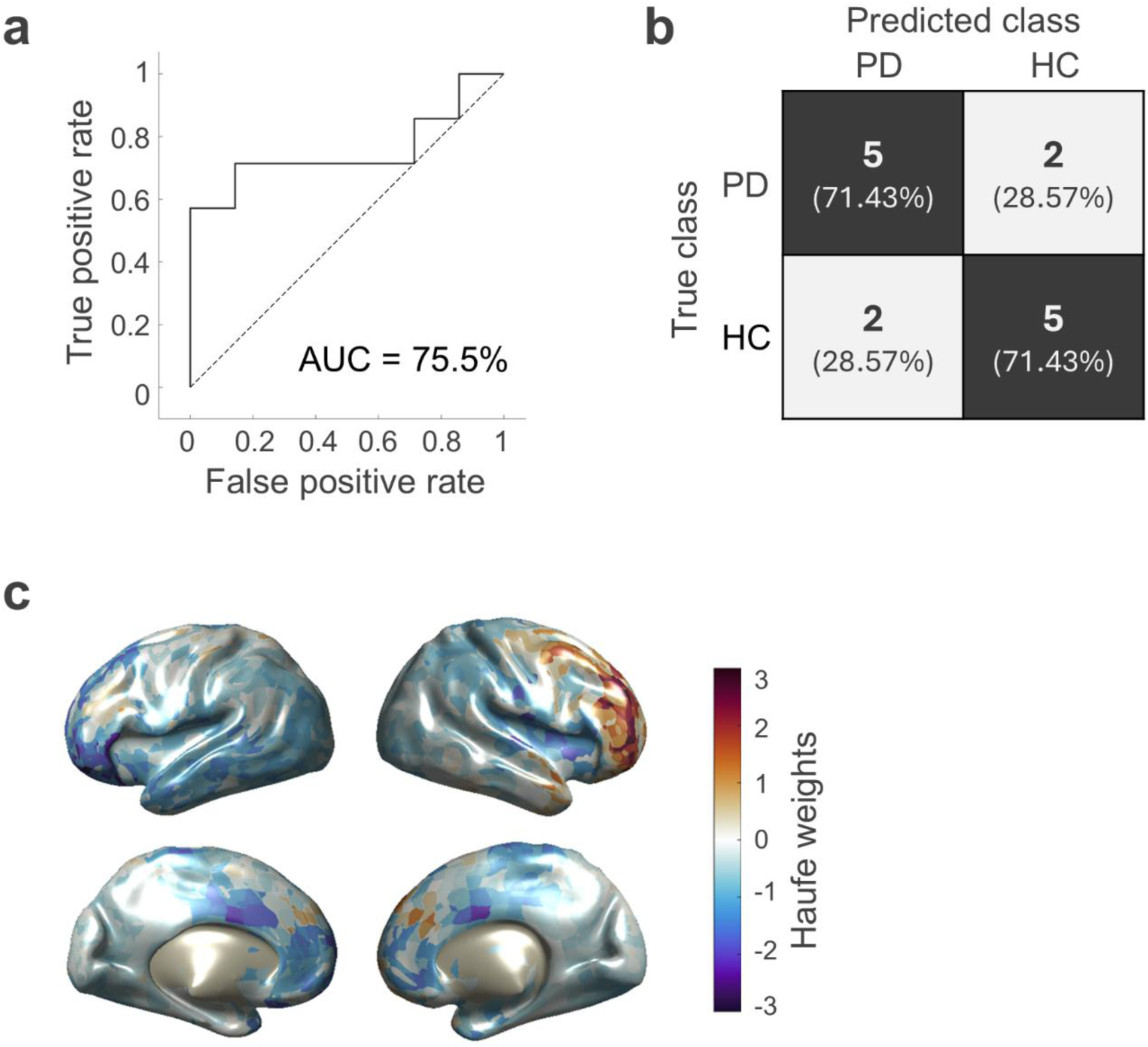
Performance of the final classification model on the hold-out test set. **(a)** Receiver operating characteristic (ROC) curves of the final SVM model on the hold-out test set (N = 14), under the no-normalization/no-PCA (1921 voxels as features) scheme. **(b)** Confusion matrix of the final model: true positives (upper-left cell), true negatives (bottom-right cell), false positives (bottom-left cell), false negatives (upper-right cell). **(c)** Haufe weights per voxel contributing to the model’s prediction on the hold-out test set. The highest positive weights drive the prediction toward the positive class (PD), whereas the most negative weights drive it toward the negative class (HC). Abbreviations: AUC: area under the curve; PD: Parkinson’s disease; HC: healthy controls.

After training the model, the contribution of each feature to the model’s decision was assessed using the Haufe transformation (Haufe et al., 2014). The resulting weights indicate the contribution of each voxel to the model’s predictions on the hold-out test set. As shown in Figure 4c, the highest positive weights, which drive predictions toward the PD class, were concentrated in right frontolateral regions and superior frontomedial areas. Conversely, the most negative weights, which drive predictions toward the HC class, were located in the left frontolateral cortex, bilateral frontomedial areas, and occipital-temporal regions.

## 4. Discussion

In this study, we investigated the potential of the brain’s natural frequencies as biomarkers for Parkinson’s disease by extracting single-subject natural frequency maps from EEG recordings and evaluating their utility for classification using machine learning. Our results provide converging evidence for disease-related alterations in intrinsic cortical oscillatory dynamics and highlight the relevance of natural frequencies as informative and interpretable features for PD detection.

Group-level patterns show a relative shift toward lower natural frequency values in PD patients compared to HC participants, particularly in frontomedial and left frontolateral cortical regions. This slowing is consistent with prior studies describing EEG/MEG abnormalities in PD as a general reduction of alpha and beta power and an increase in delta and theta power, often referred to as “EEG slowing” (Bosboom et al., 2006; Kotini et al., 2005; Morita et al., 2009; Serizawa et al., 2008; Soikkeli et al., 1991; Stoffers et al., 2007; Zhao et al., 2025). However, unlike most prior studies that focused on spectral power, we directly assessed peak frequency values, revealing that intrinsic oscillatory slowing in PD is not merely a change in amplitude but a genuine shift in oscillatory frequency.

The feature contribution analysis using the Haufe transformation (Haufe et al., 2014) further supports the functional relevance of these changes. Haufe et al. (2014) showed that, in the context of linear predictive models, the corresponding forward model—or activation pattern—can be recovered by multiplying the data covariance matrix with the model weights, which effectively represents the covariance between the features and the predicted target variable. This transformation converts the classifier’s weight vector into an interpretable activation pattern (Haufe weights), enabling a meaningful interpretation of feature contributions. In this framework, positive pattern values indicate features whose higher expression covaries positively with the model’s prediction (i.e., favoring the PD class), whereas negative values indicate features whose higher expression covaries negatively with the prediction (i.e., favoring the control class). Therefore, in our study, positive Haufe values can be interpreted as regions where higher frequency values are associated with a higher probability of being classified as PD, as for right frontolateral cortex and superior frontomedial areas. Conversely, negative Haufe values indicate regions where higher frequency values are associated with a higher probability of being classified as HC, as observed in the left frontolateral cortex, bilateral frontomedial regions, and occipital-temporal cortices. These findings suggest that slowing in PD has distinct regional signatures that can be leveraged for classification, providing a mechanistic link between altered oscillatory dynamics and disease-specific cortical patterns.

Interestingly, a hemispheric asymmetry can be observed, particularly over the lateral prefrontal regions. This asymmetry is consistent with the typical lateralized onset of PD symptoms, where motor dysfunction often begins unilaterally (Kalia & Lang, 2015). Moreover, electrophysiological findings have shown that in PD patients, frontal regions exhibit a higher degree of beta-band power lateralization compared to healthy participants (Mostile et al., 2015). In our study, higher Haufe weights were assigned to the right frontolateral cortex, whereas lower Haufe weights were found over left frontolateral and frontomedial regions. These findings support the notion of a lateralized oscillopathy in PD, in which abnormal beta and low-frequency dynamics reflect pathophysiologically meaningful hemispheric asymmetries in cortical function.

Among the three classifiers tested, SVM yielded the highest overall performance, achieving its best results when neither normalization nor PCA was applied. The comparable performance of SVM with PCA-transformed features indicates that dimensionality reduction does not improve classification accuracy in this context. This outcome aligns with previous observations that PCA is highly sensitive to data characteristics and that the variance it captures does not necessarily translate into enhanced classification performance (Janecek et al., 2008). Normalization did not improve model performance either, even though some studies reported decreased SVM performance with EEG features under no normalization schemes (Akbulut, 2020; Apicella et al., 2023). Natural frequency values are inherently within the same scale and a comparable range, which may reduce the need for additional scaling. Taken together, these findings suggest that voxel-level frequency values are sufficiently informative and that further transformations do not substantially improve performance. From a methodological perspective, this has important implications for reproducibility and interpretability. In line with Woo et al. (2017), the use of features that retain neurophysiological meaning and emerge from neuroscientifically plausible processing steps is crucial for building predictive models that are both interpretable and translationally relevant. Moreover, the spatial patterns revealed by the Haufe-transformed weights demonstrate that the classifier leverages biologically meaningful cortical regions rather than arbitrary feature combinations, thereby supporting the mechanistic validity of our approach.

The hold-out evaluation yielded an AUC of over 75% and accuracy, sensitivity, and specificity of over 71%. These results are in line with previous reports using machine learning models based on MEG/EEG-derived features for PD detection (Aljalal et al., 2022; Anjum et al., 2020; Chaturvedi et al., 2017; Lensky, 2025; Roberts et al., 2024; Vanneste et al., 2018; Waninger et al., 2020; Yuvaraj et al., 2018), where accuracies vary between 65% to an impressive 99%. While these studies have made considerable advances toward achieving a biomarker-based diagnosis of PD, they share certain methodological limitations, such as relatively small sample sizes (14–30 PD patients) (Aljalal et al., 2022; Anjum et al., 2020; Lensky, 2025; Vanneste et al., 2018; Waninger et al., 2020; Yuvaraj et al., 2018), unmatched control groups and data imbalance (Aljalal et al., 2022; Chaturvedi et al., 2017; Lensky, 2025; Roberts et al., 2024; Vanneste et al., 2018; Waninger et al., 2020), and non-standardized preprocessing protocols (Aljalal et al., 2022; Lensky, 2025; Vanneste et al., 2018; Waninger et al., 2020; Yuvaraj et al., 2018). Importantly, most rely exclusively on internal cross-validation, without additional external validation (Aljalal et al., 2022; Chaturvedi et al., 2017; Lensky, 2025; Roberts et al., 2024; Vanneste et al., 2018; Yuvaraj et al., 2018).

Here, we sought to overcome these limitations to enhance methodological strength and improve the interpretability of our results. First, this study integrated data from three independent EEG databases and employed sex- and age-matched samples of PD patients and HC, thus addressing common issues of small or imbalanced cohorts in previous research. Second, the analysis pipeline is fully standardized and reproducible, with preprocessing based on the automatic DISCOVER-EEG protocol (Gil Ávila et al., 2023) and subsequent analysis steps following openly accessible and reproducible procedures (Arana et al., 2025; Capilla et al., 2022). Third, the model was evaluated on an independent hold-out test set, providing a reliable assessment of its generalizability. Finally, by using voxel-wise natural frequencies—biologically grounded and interpretable features—together with the Haufe transformation for mapping feature contributions, the approach preserves neurophysiological meaning and enables mechanistic insights into disease-related cortical alterations. This highlights the value of combining transparent machine learning practices with neurophysiologically meaningful features for advancing translational EEG biomarkers in PD.

Nevertheless, our study has also some limitations that should be acknowledged. Although our sample size is larger than in several previous studies, it remains modest for training machine learning models. Combining three EEG datasets with different acquisition protocols enhances the generalizability of our findings but may also constraint overall model accuracy by introducing some heterogeneity. In particular, the differences in the number and arrangement of EEG channels across datasets (TDBRAIN: 26; PD LPC: 64; BrainLat: 128 channels) led us to use a common subset of 25 electrodes to standardize localization errors across datasets. Low-density EEG has been shown to increase source-localization error (Allouch et al., 2023; Song et al., 2015), which reduces the spatial precision of reconstructed cortical activity and limits the detection of subtle regional differences that could otherwise contribute to higher classification accuracy. Moreover, detailed information on disease stage and medication status was unavailable for several participants, which introduces additional heterogeneity. It is also possible that some of the control participants were not free from underlying neurodegenerative processes, as diagnostic accuracy by clinicians during the early stages of PD remains challenging (Rizzo et al., 2016). The standard clinical diagnosis is typically based on the onset of motor symptoms; however, the prediagnostic stage of PD can last for decades before the appearance of motor symptoms (Kalia & Lang, 2015; Postuma & Berg, 2016). During this period, 40–60% of dopaminergic neurons may already be lost, and synaptic function can be reduced by up to 80% before clinical signs allow a definitive diagnosis of PD (Kalia & Lang, 2015; Postuma & Berg, 2016). These neural changes are indeed accompanied by alterations in neural oscillations (Nimmrich et al., 2015; Schnitzler & Gross, 2005). Therefore, disease progression and medication effects should be carefully considered when interpreting changes in oscillatory activity in PD (Nimmrich et al., 2015; Schnitzler & Gross, 2005). Future studies using larger and more homogeneous samples, potentially incorporating longitudinal designs or applying stratification by disease stage, could further elucidate how alterations in natural frequencies evolve throughout the clinical course and thereby improve the interpretability of predictive models based on neurophysiological signatures.

Taken together, our results suggest that natural frequency maps could serve as accessible, cost-effective, and physiologically relevant biomarkers for PD, complementing clinical assessments and traditional EEG analyses based on band power. The combination of EEG-based frequency mapping with machine learning provides a framework for early detection, disease monitoring, and potentially individualized profiling, given the stability of natural frequencies at the single-subject level in healthy individuals (Arana et al., 2025). Importantly, these maps capture region-specific alterations that inform the underlying pathophysiology of PD, bridging electrophysiological measurements with clinical manifestations and providing an approach that may generalize across the broader spectrum of oscillopathies.

## 5. Conclusions

In this study, we analyzed single-subject brain natural frequency maps derived from EEG recordings to assess their potential as biomarkers for Parkinson’s disease. Our results show region-specific slowing of intrinsic cortical oscillations in PD, particularly in frontomedial and left frontolateral regions, and reveal hemispheric asymmetries consistent with the lateralized onset of clinical symptoms. Classification using voxel-wise natural frequencies achieved robust performance, indicating that these simple yet physiologically meaningful features can reliably differentiate patients from healthy controls. Altogether, these findings highlight the translational potential of natural frequency analyses, offering a promising avenue for biomarker development in oscillopathies.

## 6. Data availability

The data used this study are available in the Two Decades-Brainclinics Research Archive for Insights in Neurophysiology (TDBRAIN) database (https://www.brainclinics.com/resources; Van Dijk et al., 2022), the Linear Predictive Coding for PD at Rest (PD LPC Rest) dataset (https://predict.cs.unm.edu/downloads.php; Anjum et al., 2020), and the Latin American Brain Health Institute (BrainLat) dataset (https://www.synapse.org/Synapse:syn51549340/wiki/624187; Prado et al., 2023).

## 7. Code availability

The DISCOVER-EEG preprocessing pipeline is available in GitHub (https://github.com/crisglav/discover-eeg; Gil Ávila et al., 2023). All codes for subsequent preprocessing and data analysis are available at https://github.com/necog-UAM.

## Acknowledgements

We are thankful to all the researchers and technicians involved in the Two Decades-Brainclinics Research Archive for Insights in Neurophysiology (TDBRAIN), the Linear Predictive Coding for PD at Rest, and the Latin American Brain Health Institute (BrainLat) projects for recording and making publicly available the EEG datasets used in our study. The authors also thank Enrique Stern, Juan José Herrera-Morueco, Jorge San Segundo, and the team of the Institute for Biomagnetism and Biosignal Analysis for their valuable advice regarding the manuscript.

This work was supported by Ministerio de Ciencia e Innovación / Agencia Estatal de Investigación, Spain / FEDER/FSE+, UE (MCIN/AEI/ 10.13039/501100011033/FEDER/FSE+, UE; PID2021-125841NB-I00; PID2024-161032NB-I00 to AC); and the Comunidad de Madrid, Spain (IND2022/SOC-23652 to LA).

## 8. Author contributions

Lydia Arana: Conceptualization, Data curation, Formal analysis, Methodology, Software, Visualization, Writing – original draft preparation; Joachim Gross: Conceptualization, Methodology, Software, Resources, Writing – review & editing, Supervision; Almudena Capilla: Conceptualization, Funding acquisition, Methodology, Resources, Writing – original draft preparation, Supervision.

## Notes

### Competing Interest Statement

The authors have declared no competing interest.

